# New predictive biomarkers for plasma leakage in dengue fever

**DOI:** 10.1101/2025.07.03.25330389

**Authors:** Samaneh Moallemi, Anne Poljak, Nicodemus Tedla, Chathurani Sigera, Praveen Weeratunga, Deepika Fernando, Senaka Rajapakse, Andrew R. Lloyd, Chaturaka Rodrigo

## Abstract

Plasma leakage, which is a defining feature of severe dengue, lacks early predictive biomarkers critical for timely clinical decision-making. We aimed to characterize early biomarkers of dengue-associated plasma leakage using two complementary high-throughput proteomics platforms. Plasma samples were collected from 222 patients with dengue during their early febrile phase (stratified by subsequent development of plasma leakage and prior dengue exposure), 50 non-dengue patients with febrile illnesses, and 6 healthy controls. Proteomic profiling of pooled plasma was performed using liquid chromatography–tandem mass spectrometry (LC-MS/MS) and SomaScan aptamer-based platforms. Differential protein expression, and pathway analyses were conducted to identify early biomarkers of plasma leakage. We identified 23 differentially expressed proteins detected across both platforms that distinguished patients who subsequently did or did not develop plasma leakage. Fourteen of these proteins are highly expressed in the liver, demonstrating a central role of early hepatic dysfunction in plasma leakage. Functional analysis of these biomarkers revealed convergent roles in endothelial dysfunction, immune dysregulation, extracellular matrix remodelling, and metabolic reprogramming. Proteomic screening using two powerful complementary methods applied to pooled plasma samples identified 23 biomarkers associated with subsequent plasma leakage. These biomarkers must be prioritized for validation in other cohorts to demonstrate generalisability.

## Introduction

Plasma leakage is a defining pathophysiological hallmark of severe dengue but is only seen in 35-38% of patients with symptomatic dengue fever[1]. While not all patients with plasma leakage develop shock or organ failure, nearly all severely ill dengue patients exhibit preceding plasma leakage, making it a biologically-determined, clinically-actionable and objectively measurable intermediate outcome[2]. In contrast to the broader World Health Organization (WHO) severity classifications like severe dengue and dengue haemorrhagic fever, which are composite clinical criteria often inconsistently applied[3], plasma leakage is an objectively demonstrable single criterion diagnosed by serial haemoconcentration or ultrasonography[1].

Currently, the risk of plasma leakage cannot be predicted during the “early” febrile phase of dengue[4], when decisions regarding hospital admission, intensive monitoring and supportive care must be made, to avoid complications. This uncertainty contributes to both unnecessary hospitalisations and missed opportunities for timely intervention—particularly in resource-limited settings where seasonal surges in dengue incidence strain clinical capacity[5]. The timing of plasma leakage (typically days 5-7 post onset of fever), which follows peak viraemia, supports a model in which host immune responses, rather than direct viral cytopathy, drive endothelial dysfunction. This is especially evident in secondary infections, where antibody-dependent enhancement amplifies immune activation and contributes to vascular inflammation[6].

A limited number of proteomic studies, along with several targeted investigations measuring individual proteins, have implicated mechanisms such as cytokine release, endothelial activation, and glycocalyx degradation in plasma leakage[7–15] . However, these findings remain fragmentary, not specific to plasma leakage, and relied on a single proteomics platform without cross-validation. To explore early host responses preceding plasma leakage, we conducted what is, to our knowledge, the first study to apply a high-throughput, dual-platform proteomic discovery strategy. By integrating two complementary technologies, we aimed to achieve broad and sensitive proteome coverage across key biological pathways to identify predictive biomarkers for dengue plasma leakage.

## Methods

### Cohort characteristics and clinical assessment

Stored plasma and clinical data were obtained from the Colombo Dengue Study (CDS), a prospective observational cohort conducted in a collaboration between the University of Colombo, Sri Lanka, and University of New South Wales, Australia. Participants with suspected dengue were recruited between 2017-2020 at the National Hospital of Sri Lanka within 96 hours of fever onset, according to the WHO clinical case definition. A comprehensive description of the study methodology of CDS has been published previously[16].

Dengue infection was confirmed by bedside detection of non-structural protein 1 (NS1) antigen (SD Bioline, Alere SD, USA) and RT-qPCR (SuperScript III Platinum One-Step Quantitative RT-PCR System, Invitrogen, USA). Prior dengue exposure (primary vs. secondary dengue) was determined by using an anti-dengue envelope IgG assay (Euroimmun, Germany), with IgG presence at early infection being indicative of prior infection. Clinical management was provided based on local guidelines, independent of these test results.

Participants underwent a daily clinical and laboratory evaluation. Plasma leakage was defined by standardised criteria: ≥20% haematocrit elevation from baseline, absolute haematocrit≥45%, or ultrasonographic evidence of third-space fluid accumulation (pleural effusions or ascites). No participants exhibited plasma leakage at enrolment.

Ethical approvals were granted by the University of Colombo (EC/17/080), UNSW Sydney (HC220706), and St Vincent’s Hospital Sydney (for healthy controls) (HREC/13/SVH/145). Written informed consent was obtained from all participants. All research were performed in accordance with the principles of Declaration of Helsinki.

### Study design

Plasma from confirmed dengue patients were stratified into two main groups of equal size with and without plasma leakage for comparison (PL+ vs. PL-). These were subdivided further into four clinical categories: primary dengue without plasma leakage (PD+PL-), primary dengue with plasma leakage (PD+PL+), secondary dengue without plasma leakage (PD-PL-), and secondary dengue with plasma leakage (PD-PL+). Age- and sex-matched pools were generated with 50 individuals per pool, and one pool per category. Pool size was informed by power calculations from our previous study[17](∼51 individuals per group); 50 individuals per pool were selected to maintain statistical power while minimising inter-individual variability and preserving representative heterogeneity. All participants were free of known chronic medical conditions such as diabetes, cardiovascular disease, chronic kidney and liver disease, to minimise confounding effects in the proteomic analysis. An additional 22 severe dengue cases (all secondary dengue with plasma leakage), 50 non-dengue fever patients, and 6 healthy controls (of Sri Lankan ethnicity recruited in Australia) were also processed as three additional categories. Altogether, plasma from 278 patients (222 dengue patients, 50 non-dengue fever patients, 6 healthy controls) were analysed. The number of individuals per pool was arbitrarily capped at 50, except for the single pool of 22 individuals which included all severe dengue patients from CDS. Each of the 7 plasma pools were analysed in two technical replicates. The data and analysis presented in this paper is limited to identifying predictive biomarkers for plasma leakage that were verified across both proteomic platforms used as described below.

### Plasma proteomic analyses

Two complementary proteomic platforms were used: liquid chromatography–tandem mass spectrometry (LC-MS/MS) and SomaScan aptamer-based proteomic profiling. LC-MS/MS analysis was performed with high-abundance protein depletion and technical duplicates.

SomaScan aptamer-based analysis targeting ∼11,000 proteins were performed by SomaLogic (Boulder, CO, USA) with adaptive normalization by maximum likelihood. Further details of proteomics workflow, and computational analyses are provided in the Supplementary Material.

### Statistical analysis

For data from both platforms, differential expression analysis was conducted using the limma package (R version 4.2.2), applying linear modelling and empirical Bayes moderation.

Presence-absence filtering was used to retain proteins consistently detected across all biological samples within each pooled group. Log2-transformation, technical replicate correlation, sample quality weighting, and robust variance modelling were applied. Group comparisons focused on plasma leakage versus no leakage across all dengue cases, followed by separate analyses within primary and secondary dengue infections.

For LC-MS/MS data, proteins with a log[fold change ≥1 and p ≤ 0.05 were considered to be significantly differentially expressed. For SomaScan, a threshold of log[fold change ≥0.2 and p ≤0.05 was applied. As previously recommended[18], this more permissive threshold improves sensitivity given the non-linear signal response of some aptamers at high protein concentrations. Only proteins consistently up- or down-regulated across both platforms were considered for analysis in this paper, as these represent the most robust cross-validated biomarkers. Spearman correlations were used to assess concordance between platforms.

Expression patterns of cross-validated proteins from the overall analysis, including fold-change values and regulatory direction, were also compared between primary and secondary dengue infections as a subgroup analysis. The Human Protein Atlas was consulted to assess tissue-specific expression and subcellular localisation. Statistical analyses and visualizations were performed with R (v4.2.2) using built-in functions and the packages limma, ggplot2, and corrplot.

## Results

Demographic comparisons showed no statistically significant differences in mean age or the proportion of males / females in patients with and without plasma leakage (Supplementary Tables 1, 2). In total, 1,358 proteins were detected by LC-MS/MS (Table S3) and 9,665 unique protein targets by SomaScan (Table S4), after excluding multiple measurements per protein across all groups. Of these, a total of 910 proteins across all samples were detected by both platforms (Table S5). Technical reproducibility metrics for both methods are presented in the Supplementary Data (Figures S1, S3, S4).

### Proteome signatures associated with dengue-associated plasma leakage

In the main analysis which compared between groups with and without plasma leakage (PL+ vs. PL-) without stratifying by prior dengue exposure, LC-MS/MS analysis revealed 108 differentially expressed proteins (DEPs) using both differential expression (log[fold change ≥1, p≤0·05) and presence/absence analyses, with 88 upregulated and 20 downregulated in PL+ patients (Figure 1A, Table S6), including 7 proteins detected exclusively in PL+ samples and 2 exclusively in PL– samples (Figure 1C). SomaScan profiling identified 3,181 DEPs (log[fold change ≥0·2, p≤0·05), comprising 1,634 upregulated and 1,547 downregulated proteins in PL+ patients (Figure 1B, Table S7). Between-platform comparisons revealed that 23 DEPs were identified by both platforms and hence cross-validated as early biomarkers of plasma leakage (Table 1).

**Figure 1.**
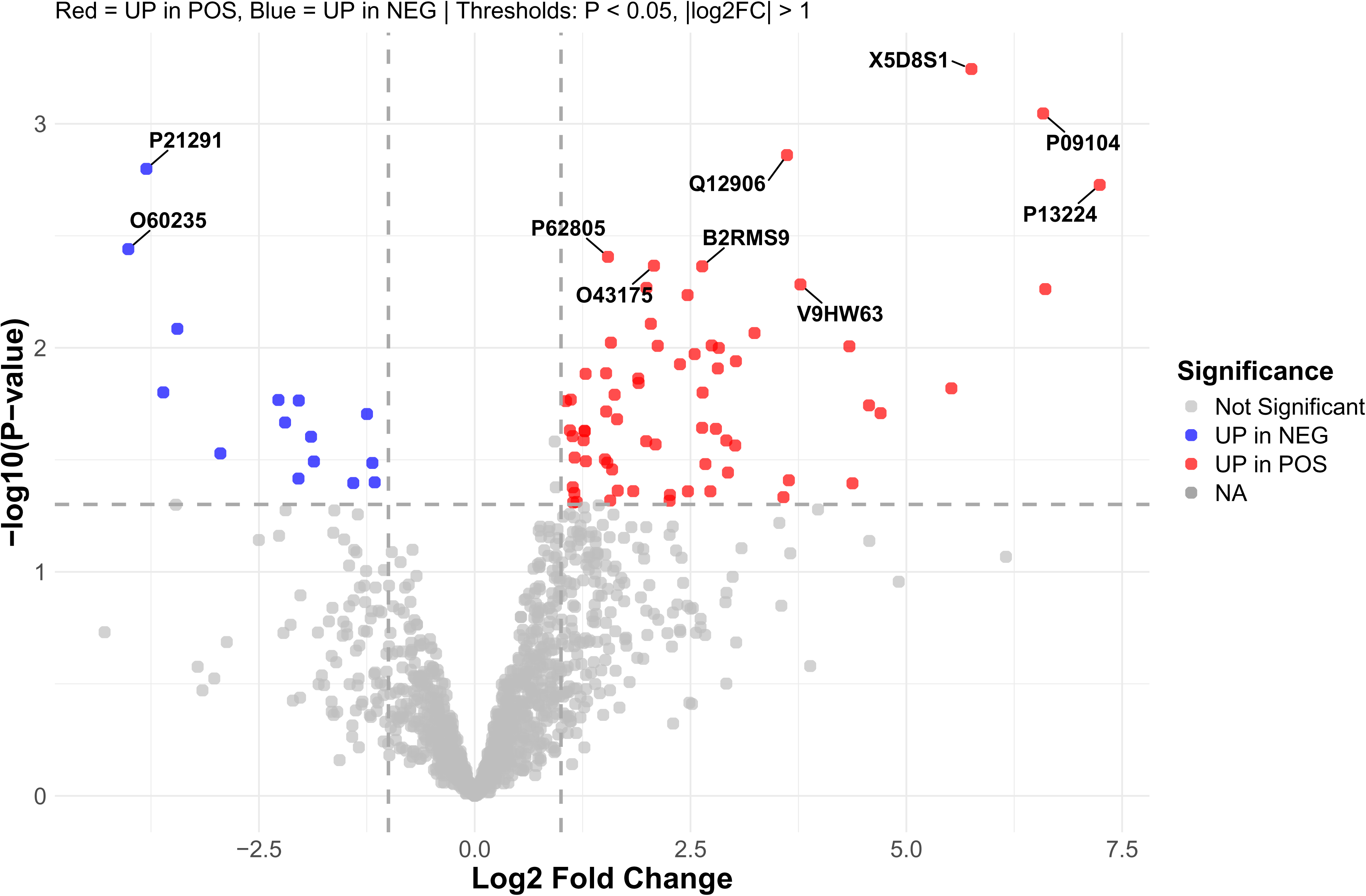
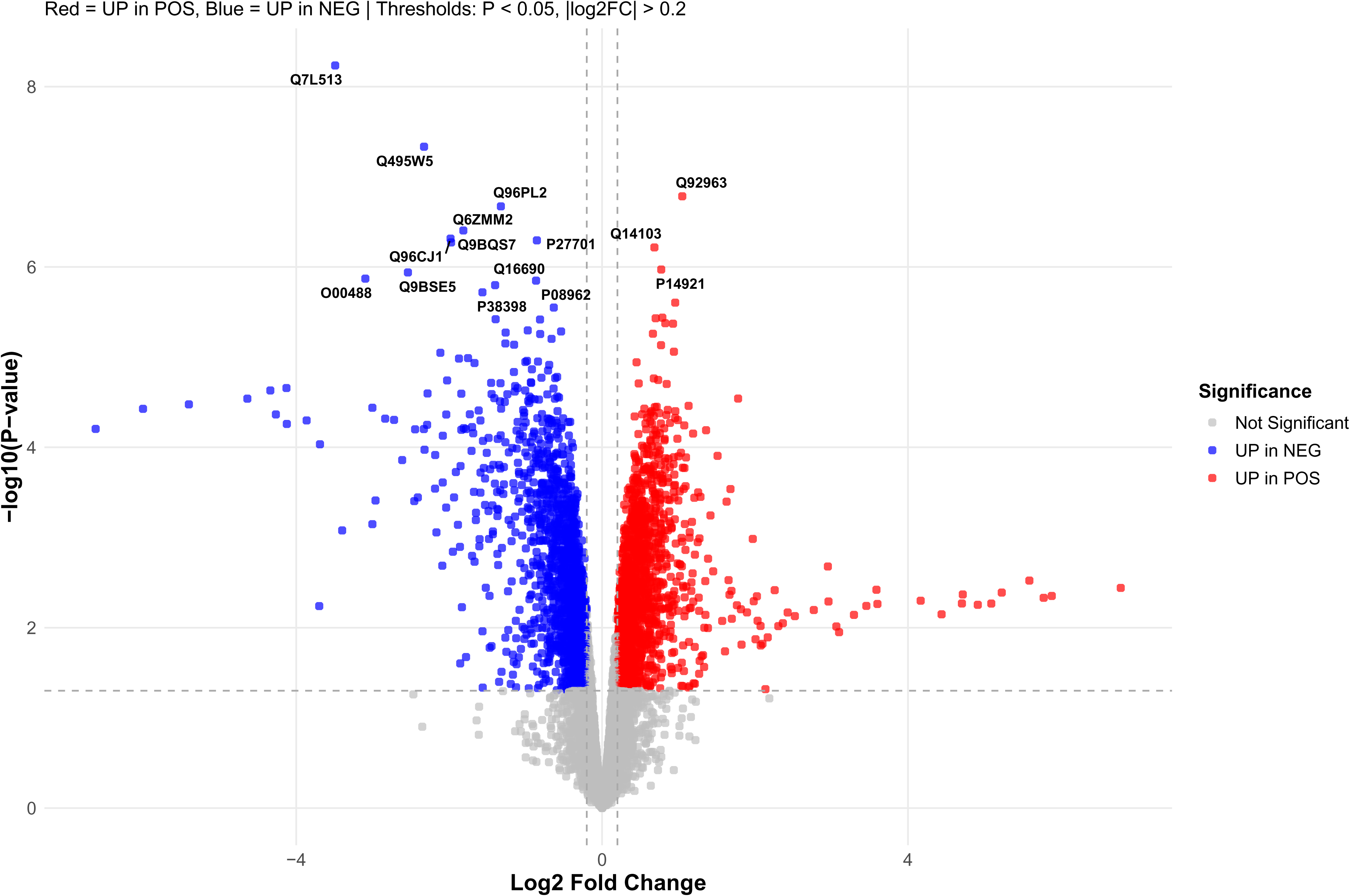
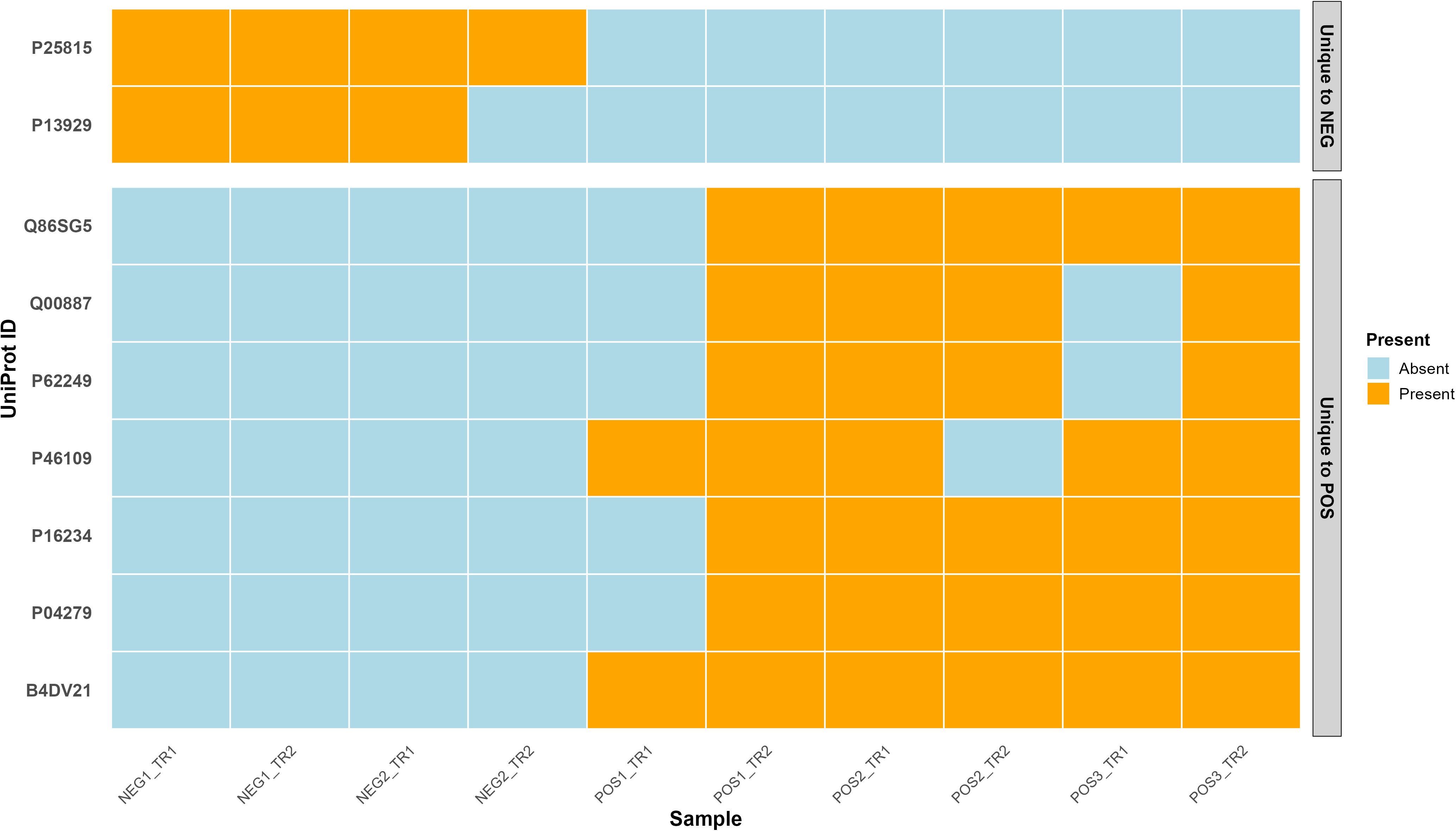
Differential protein expression in febrile-phase plasma from dengue patients who subsequently developed plasma leakage (POS) versus those who did not (NEG). Proteins are identified by their UniProt ID in all panels. (A) Volcano plot showing differentially expressed proteins detected by LC-MS/MS (log□ fold change ≥1, p ≤ 0.05). A total of 107 proteins were significantly differentially expressed, including 87 upregulated (red) and 20 downregulated (blue). (B) Volcano plot of SomaScan data (log□ fold change ≥0.2, p ≤ 0.05), identifying 3,181 DEPs: 1,634 upregulated and 1,547 downregulated proteins. Selected annotated proteins are highlighted based on statistical significance and fold change. (C) Heatmap showing proteins consistently detected in all biological samples (including technical replicates) of one group (PL+ or PL–) but absent in the other. (Y-axis displays UniProt IDs, corresponding to the following proteins: S100P (P25815), Beta-enolase (P13929), S100-A7A (Q86SG5), Pregnancy-specific beta-1-glycoprotein 9 (Q00887), Small ribosomal subunit protein uS9 (P62249), Crk-like protein (P46109), Platelet-derived growth factor receptor alpha (P16234), Semenogelin-1 (P04279), and a protein similar to Dimethylglycine dehydrogenase (B4DV21).) Each column represents a technical replicate (TR1, TR2) of a biological sample; “POS” indicates PL+ samples and “NEG” indicates PL– samples. The number after POS or NEG shows the sample stratification by prior dengue exposure (POS1 – primary dengue with plasma leakage, POS2 – secondary dengue with plasma leakage, POS3 – severe dengue; NEG1 – primary dengue without plasma leakage, and NEG2 – secondary dengue without plasma leakage). Orange cells represent protein presence; white cells indicate absence. Two proteins (P25815, P13929) were uniquely detected in PL–samples, while seven proteins were uniquely present in PL+ samples.

**Table 1.**
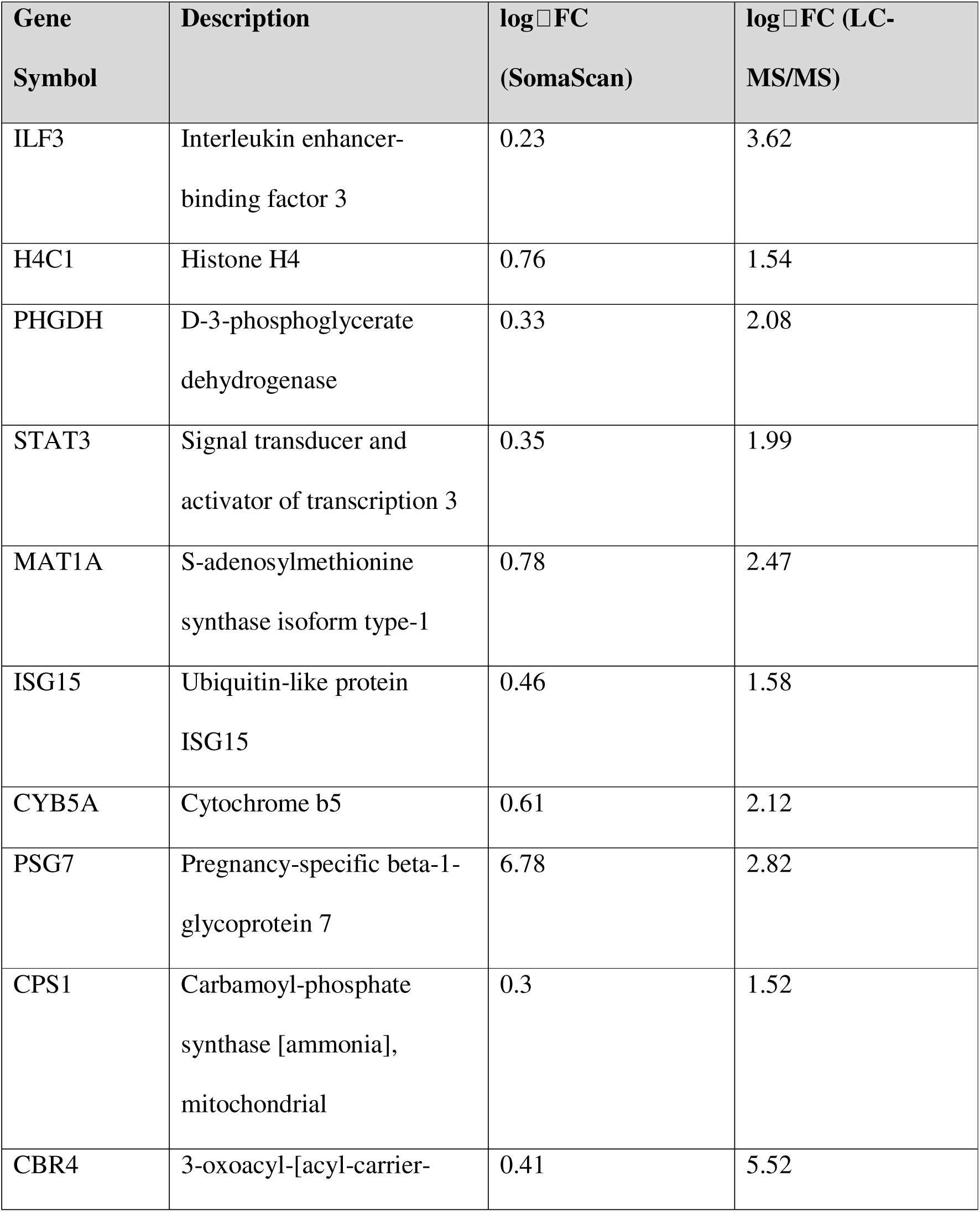
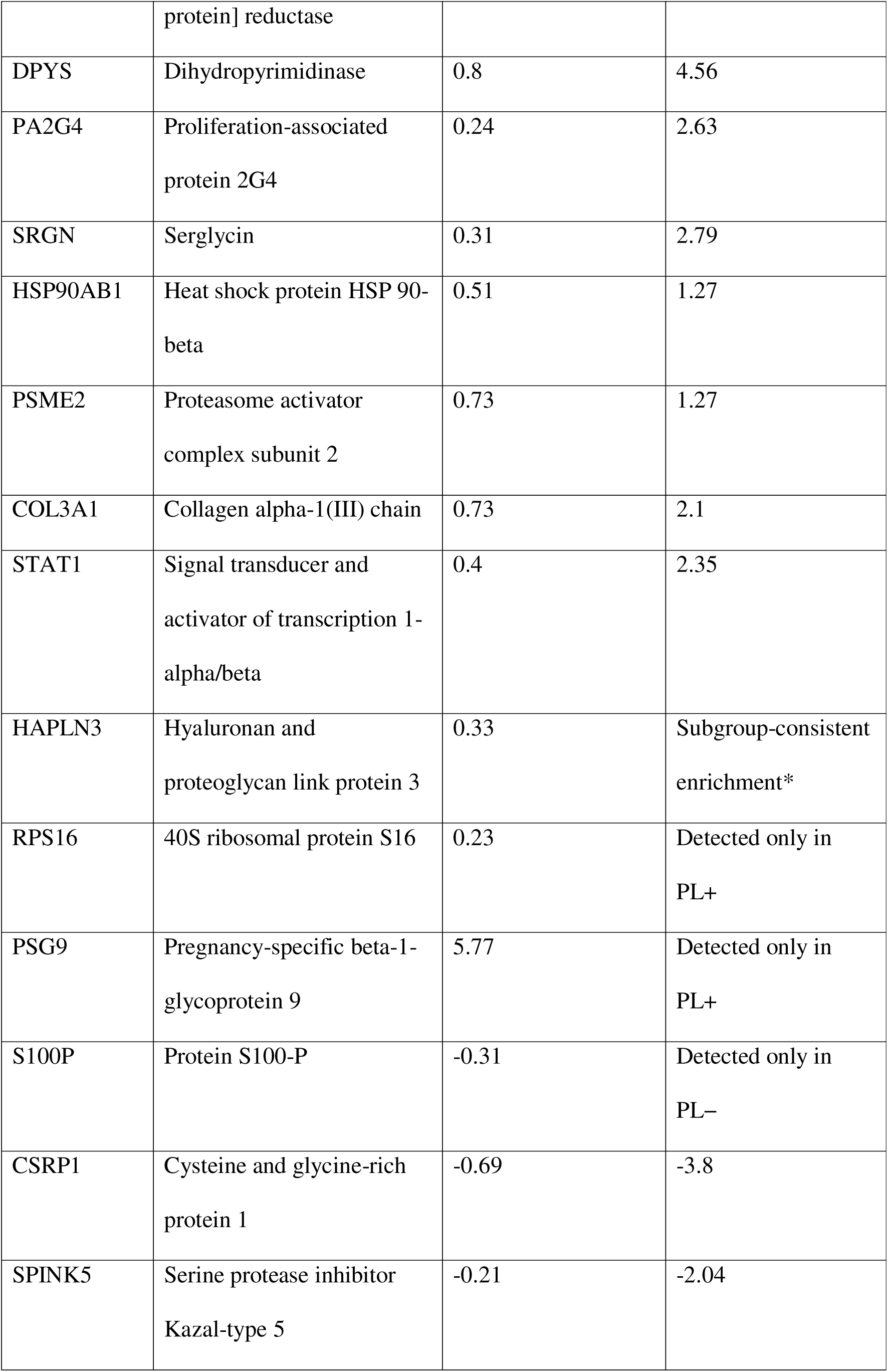
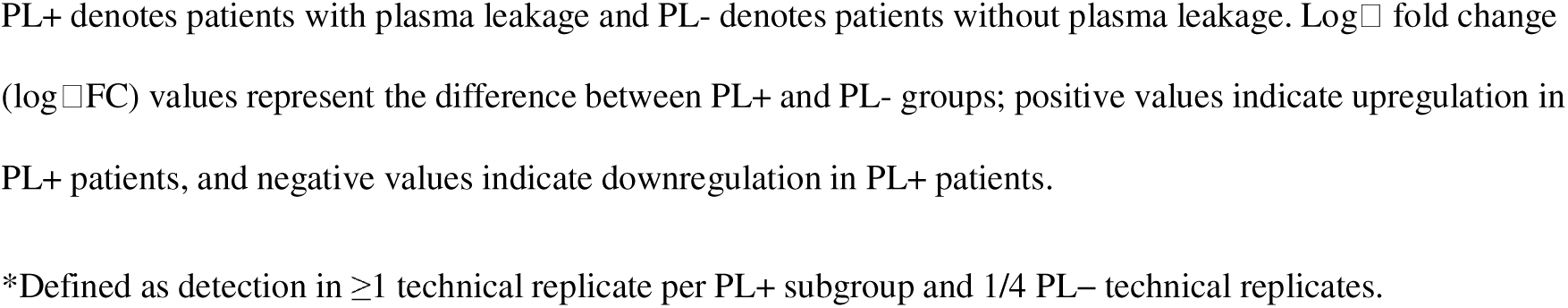
The 23 biomarkers associated with plasma leakage in dengue and cross validated in both proteome analysis platforms.

Throughout this manuscript, biomarkers described as “identified” refer to those meeting the predefined statistical significance and fold-change criteria outlined in the Methods.

### Subgroup analysis by infection history

In primary dengue samples out of the 23 biomarkers identified in the main analysis, six (COL3A1, MAT1A, ILF3, PHGDH, STAT3, ISG15) were detected by SomaScan and three (MAT1A, CSRP1, PSG7) were detected by LC-MS/MS respectively. The only biomarker (MAT1A) detected across both platforms did not show an association in the same direction between the two platforms.

In secondary dengue samples, out of the 23 biomarkers identified in the main analysis. fifteen (ISG15, CBR4, COL3A1, CPS1, DPYS, H4C1, HSPA1A, MAT1A, PSME2, RPS16, SRGN, STAT1, STAT3, HSP90AB1, SPINK5) were detected by SomaScan and eleven (SRGN, ILF3, DPYS, STAT3, CBR4, CYB5A, ISG15, H4C1, PHGDH, HSP90AB1, PA2G4) were detected by LC-MS/MS. Of these, (ISG15, SRGN, STAT3, RPS16, H4C1, DPYS, CBR4) were consistently detected across both platforms in the same direction. Five biomarkers (ISG15, STAT3, COL3A1, ILF3, PHGDH) were detected in both primary and secondary dengue patients by at least one platform.

In addition, ten proteins in primary dengue and fifteen in secondary dengue were significant in the stratified analyses but were not significant in the combined cohort analysis and were therefore not included among the 23 biomarkers reported in the main analysis (Tables S15 and S18).

Functional analysis of biomarkers detected by at least one platform revealed that both primary and secondary dengue shared biomarkers involved in interferon signalling (ISG15, STAT3), extracellular matrix formation (COL3A1), metabolic processes (MAT1A, PHGDH), and RNA binding (ILF3). Secondary dengue patients had additional biomarkers related to immune signalling (STAT1), cellular stress responses (HSPA1A), protein degradation (PSME2), and ribosomal function (RPS16), while primary dengue patients had biomarkers of cytoskeletal regulation (CSRP1) and immune modulation (PSG7).

## Discussion

This high throughput proteomic study leveraged two orthogonal platforms, mass spectrometry and aptamer-based proteomics, to identify 23 consistently differentially expressed, cross-validated early biomarkers of dengue patients with and without plasma leakage. Collectively they suggest early disruption of immune and metabolic pathways and structural integrity of the vascular endothelium by increased activation of cytokine and stress response pathways via key signalling mediators (STAT1[19], STAT3[19],ISG15[20], and HSP90AB1[21]) by mediators of heightened proteasomal activity (PSME2[22]).

Our findings advance previous proteomic studies of dengue, which have typically focused on general infection markers rather than leakage-specific signatures[7–15]. For example, Choudhury et al[12]. highlighted broad inflammatory and endothelial alterations in the proteome, but did not stratify findings by plasma leakage status, limiting insights into the molecular events underlying vascular dysfunction. Several proteins found to be upregulated in the plasma leakage group in this study have been previously implicated in dengue pathogenesis, although not all in the context of severe disease. Even when they had been associated with severe disease (e.g., (STAT1, STAT3, S100P, and COL3A1), none had been flagged as early predictors of that outcome[19,23,24]. Previous proteomic studies identified STAT1 and STAT3 as central regulators of dengue shock syndrome (DSS) via IFN and NF-κB signaling[19]. STAT3 also promotes viral replication, and its inhibition reduces dengue virus protein expression[23]. HSP90AB1, another upregulated protein, facilitates dengue virus entry via lipid rafts in monocytes and epithelial cells, although its role in severe clinical outcomes is unclear[21]. COL3A1, an extracellular matrix protein, has been associated with immune modulation and vascular remodelling in dengue, including interactions with endogenous retroviral elements linked to increased endothelial permeability[24]. HAPLN3, which facilitates extracellular matrix binding and matrix stabilization, was previously noted to be upregulated in intermediate phenotype monocytes from dengue patients[25].

Upregulation of extracellular histone H4, linked to neutrophil extracellular traps (NETs), supports emerging evidence for the role of NET in dengue vasculopathy[26]. In contrast, S100P, which was downregulated in the plasma leakage group, has previously been reported as upregulated during DSS[19], suggesting context-dependent expression based on disease severity. Early downregulation of SPINK5, and CSRP1 alongside S100P which together are involved in vascular tone and cytoskeletal dynamics, may indicate early impaired endothelial homeostasis in patients who subsequently develop leakage. While most prior studies were focused on general dengue infection or early immune responses, our findings place these proteins within the specific context of plasma leakage, a critical knowledge gap in the field. Together, these findings support a model in which plasma leakage is driven by early inflammatory and metabolic changes, accompanied by selective disruption of endothelial structure and function. Key protein biomarkers such as STAT3, HSP90AB1, COL3A1, PHGDH, and H4C1, all offer mechanistic insight and also represent promising candidates for early prediction of vascular leakage in dengue. Notably, 14 of the 23 cross-validated proteins are highly expressed in the liver. Building on previous work which identified liver-derived biomarkers such as CRP and AST as early predictors of severe outcome[27], this analysis further expands the list of hepato-centric biomarkers associated with adverse dengue outcomes. Several metabolic regulators identified here—CPS1, a liver-derived urea cycle enzyme promoting anti-inflammatory M2 macrophage polarisation, MAT1A, which regulates methionine metabolism and interferon-mediated antiviral responses, and PHGDH, a serine biosynthesis enzyme modulating antiviral innate immunity, have not previously been linked to dengue, and represent novel candidates for further investigation.

The subgroup analysis showed a wider overlap of findings with the main analysis for secondary dengue patients than for primary dengue patients. However, five proteins (ISG15, STAT3, COL3A1, ILF3, and PHGDH) were consistently detected across both primary and secondary dengue patients by at least one platform, suggesting their potential as universal plasma leakage predictors regardless of prior exposure history, an important consideration given that most previous biomarker studies have not distinguished between primary and secondary infections. In this regard, ISG15 and STAT3 were the strongest candidates as they were detected by both platforms in secondary dengue patients and by one platform (SomaScan) in primary dengue patients. Identifying predictive biomarkers for plasma leakage without considering prior dengue exposure is also clinically more relevant because prior immune history is often not tested when managing dengue patients because such testing is expensive and difficult to interpret specially for late admissions (i.e., anti-dengue IgG may be present due to both past and current infections). Self-report by patients of their past dengue infections is also unreliable[28] given the widespread practice of presumptive treatment without diagnostic confirmation in resource limited settings, and the many differential diagnoses with similar symptoms. However, from a disease pathogenesis perspective, the stratified (subgroup) analysis is more important to identify the metabolic pathways involved after adjusting for the confounding effect of antibody dependent enhancement. An exploration of dengue pathogenesis considering the whole proteome detected by each of the platforms is beyond the scope of this paper.

This analysis considered equal numbers of primary and secondary dengue patients matched for age and sex to limit their confounding effects on proteome analysis. This is a methodological strength as well as a limitation. The latter is because the main analysis has an equal mix of primary and secondary dengue patients whereas in real-life this ratio will be weighted towards secondary dengue dominance in adult populations in endemic countries or towards primary dengue in young children, travellers from non-endemic countries or in populations where dengue is a newly emerging infection, affecting the generalizability of the results. Pooling of samples is another limitation which may have masked inter-individual variability, particularly for low-abundance proteins. The sampling window, restricted to the first 96 hours of illness, was treated as a homogeneous phase, but daily or hourly fluctuations in protein expression may have been missed due to the lack of serial sampling. Although the dual-platform design enhanced discovery power and confidence, the limited overlap between platforms was a constraint. Furthermore, given the semi-quantitative output, this study is best described as high throughput screening, and the top DEPs for each comparison should be quantitatively validated in independent cohorts to confirm their predictive utility in future.

## Conclusion

This study identified 23 early biomarkers for dengue associated plasma leakage via an extensive dual-platform proteomic screen providing insights into early disruption of immune, metabolic, and structural homeostatic pathways. A majority of these biomarkers are highly expressed in the liver indicating potential early hepatic dysfunction in patients with plasma leakage. Prioritising these biomarkers for future validation may improve early risk stratification and support the development of point-of-care prognostics for dengue.

## Data availability statement

Processed proteomic data supporting the findings of this study are provided in the supplementary Excel file available at Harvard Dataverse: https://doi.org/10.7910/DVN/HKHVSN. Requests for additional data access should be directed to the corresponding author and will be considered on a case-by-case basis following appropriate ethical and institutional approvals.

## Data Availability

The minimum dataset for this manuscript is available within the manuscript and supplementary material. Processed proteomic data supporting the findings of this study are provided in the supplementary Excel file available at Harvard Dataverse: https://doi.org/10.7910/DVN/HKHVSN. The codebook of datafields and a list of data tables is provided in the supplementary material. Requests for additional data access should be directed to the corresponding author and will be considered on a case-by-case basis following appropriate ethical and institutional approvals.

