## Supplementary Tables for "New predictive biomarkers for plasma leakage in dengue fever"

### Supplementary Table 1. Baseline characteristics of study groups

| Category | Group | N | Mean Age (years) | Male, n (%) | p-value (Age) | p-value (Sex) |
| --- | --- | --- | --- | --- | --- | --- |
| Primary dengue | Without plasma leakage (PD+PL−) | 50 | 29.34 | 40 (80.0) | 0.45 | 1.00 |
|  | With plasma leakage (PD+PL+) | 50 | 27.82 | 39 (78.0) |  |  |
| Secondary dengue | Without plasma leakage (PD−PL−) | 50 | 35.5 | 25 (50.0) | 0.58 | 1.00 |
|  | With plasma leakage (PD−PL+) | 50 | 36.6 | 24 (48.0) |  |  |
| Controls | Non-dengue febrile | 50 | 35.1 | 25 (50.0) | — | — |
|  | Healthy controls | 6 | 34.0 | 4 (66.7) | — | — |

Values are presented as mean or number (percentage). P-values compare patients with versus without plasma leakage within primary and secondary dengue groups only. Age was compared using Student’s t-test and sex using the chi-square test. No statistical comparisons were performed within the control group. P-values are approximate as analyses were based on summary statistics.

### Supplementary Table 2. Comparison of age and sex among all patients with plasma leakage and controls

| Group | N | Mean Age (years) | Male, n (%) |
| --- | --- | --- | --- |
| Patients with plasma leakage | 100 | 32.21 | 63 (63.0) |
| Patients without plasma leakage | 100 | 32.42 | 65 (65.0) |
| Controls (combined) | 56 | 34.98 | 29 (51.8) |

Age was compared across groups using one-way ANOVA (p ≈ 0.08) and sex distribution using the chi-square test (p ≈ 0.20). There were no statistically significant differences between the groups.
